# Uptake of Covid-19 vaccines among frontline workers in California state prisons

**DOI:** 10.1101/2021.11.18.21266535

**Authors:** Lea Prince, Elizabeth Long, David M. Studdert, David Leidner, Elizabeth T. Chin, Jason R. Andrews, Joshua A. Salomon, Jeremy D. Goldhaber-Fiebert

## Abstract

**Background:** Prisons are high-risk environments for Covid-19. Vaccination levels among prison staff remain troublingly low - lower than levels among residents and members of the surrounding community. The situation is troubling because prison staff are a key vector for Covid-19 transmission.

**Objective:** To assess patterns and timing of staff vaccination in California state prisons and identify individual- and community-level factors associated with being unvaccinated.

**Design:** We calculated fractions of prison staff and incarcerated residents in California state prisons who remained unvaccinated. Adjusted analyses identified demographic, community, and peer factors associated with vaccination uptake among staff.

**Setting:** California Department of Corrections and Rehabilitation prisons.

**Participants:** Custody and healthcare staff who worked in direct contact with residents.

**Main Outcomes and Measures:** Remaining unvaccinated through June 30, 2021.

**Results:** A total of 26% of custody staff and 52% of healthcare staff took ≥1 dose in the first two months of vaccine offer; uptake stagnated thereafter. By June 30, 2021, 61% of custody and 37% of healthcare staff remained unvaccinated. Remaining unvaccinated was positively associated with younger age, prior Covid-19, residing in a community with relatively low vaccination rates, and sharing shifts with co-workers who had relatively low vaccination rates.

**Conclusions and Relevance:** Vaccine uptake among prison staff in California in regular contact with incarcerated residents has plateaued at levels that pose ongoing risks—both of further outbreaks in the prisons and transmission into surrounding communities. Staff decisions to forego vaccination appear to be complex and multifactorial. Achieving safe levels of vaccine protection among frontline staff may necessitate requiring vaccination as condition of employment.

## Introduction

Prisons and jails are particularly high-risk environments for Covid-19. Since the pandemic began, there have been more than 550,000 Covid-19 cases among residents and staff in carceral settings in the United States, nearly 3,000 deaths, and infection rates among residents are 4-5 times higher than those in the general community.(1-5) In California state prisons alone, approximately one in four residents have had Covid-19 and 241 have died.(6, 7) Correctional staff are likely to be a significant source of introduction of SARS-CoV-2 infection into prisons. (4, 8)

Beginning in December 2020, California prioritized prison residents and employees for Covid-19 vaccination. Nine months later, three-quarters of residents had elected to receive at least one dose, but only 60% of employees working with direct physical access to the residents had done so. Nationally, Covid-19 vaccine coverage rates among prison staff have been lower than rates in the wider community. To redress this situation, the federal government and several states, including California, have introduced mandates, prompting lawsuits and staff resignations.(9-11)

Vaccine hesitancy among workers in high-risk settings, like prisons, is not well understood. We analyzed uptake of Covid-19 vaccines among more than 31,000 staff in California’s state prisons. The study goal was to describe patterns of vaccination in this population and identify individual and workplace characteristics associated with low vaccine uptake.

## Methods

### Data

The California Department of Corrections and Rehabilitation (CDCR) provided anonymized person-day level data. The data included comprehensive information on PCR and antigen testing and vaccination for both prison staff and residents (January 1, 2020 through June 30, 2021), plus additional information on incident cases among residents through September 25, 2021. The staff data also included information on demographic characteristics, zip code of home residence, and which shifts each staff member worked at which prison. Statistics on SARS-CoV-2 infection by zip codes came from California Department of Public Health (CDPH) data. Stanford’s institutional review board approved the study (protocol numbers IRB-55835, IRB-55671).

### Study Sample

The study focused on correctional staff who worked in CDCR prisons between the date vaccines were first offered to staff (December 22, 2020) through June 30, 2021. Thus, the study period ended before proposals for and debate over mandatory vaccination for prison staff flared in California; this period also predated the Delta variant surge (see Appendix for more detail). We restricted the analytic sample to staff who worked 5 or more shifts between April 1, 2021 (by which time vaccines were available to any staff member who wanted one) and June 30, 2021, were employed with a designation of custody or healthcare (excluding contract employees), and worked in roles that involved regular direct contact with residents (“direct care”). Two of CDCR’s 35 prisons were excluded from the analysis due to missing or incomplete staff data at those prisons. Staff with missing values for variables of interest (<1% of those who met the above eligibility criteria) were dropped from the analysis.

### Variables

The outcomes of interest in our primary analyses were indicator variables denoting staff and residents who had not been vaccinated by the end of the study period. In secondary analyses, we also considered indicators denoting staff and residents who were neither vaccinated nor had history of SARS-CoV-2 infection.

Being unvaccinated was defined as having received no doses of a Covid-19 vaccine. No history of SARS-CoV-2 infection was defined as having no confirmed positive result from a PCR or antigen test. CDCR’s staff testing program has involved a combination of mandatory and voluntary testing; most of the testing has been routinely performed by CDCR itself, although early on, some results from staff tests obtained elsewhere were included. CDCR’s resident testing program consists of routine risk-based and surveillance testing and testing in response to detected outbreaks. (12, 13)

The demographic information on staff members included age group, racial or ethnic group, gender, recorded zip code of residence, and whether the individual had any history of SARS-CoV-2 infection.

To explore the relationship between staff members’ vaccination status and two potential environmental influences—lack of vaccine uptake in communities in which they lived and among fellow workers—we created two additional variables. First, we used the CDPH area-level data to calculate the cumulative fraction of unvaccinated individuals aged 20-64 years old on June 1, 2021 in the most recent zip code in which each staff member resided (see Appendix Table 1 for counts of included zip codes by prison). Second, we calculated the proportion of unvaccinated co-workers who worked in the same job classification (custody or healthcare) on the same shift, day, and prison as the staff member being analyzed. This variable was a composite, cross-sectional measure created by assigning a weight to each staff person’s co-workers based on the total number of shifts worked together during the study period while either vaccinated (received at least 1 dose of Covid-19 vaccine) or unvaccinated (additional details in Supplementary Appendix).

### Statistical analysis

We plotted vaccine uptake and detected prior infection among custody and healthcare staff (separately) over the study period. Next, for each prison, we compared proportions of staff and residents who remained unvaccinated at the end of the study period, and then examined the cumulative risk of detected Covid-19 among residents in those prisons in the 3 months following the study period (July 1, 2021 through September 25, 2021). We repeated this approach, comparing proportions and examining cumulative risk, after redefining the “vulnerable” groups of interest to include staff and residents who were neither vaccinated nor had history of prior detected infection by the end of the study period.

Finally, to identify characteristics of staff members who remained unvaccinated, we fit staff-level multivariable probit regression models (one each for custody and healthcare staff). The outcome variable in these models was remaining unvaccinated at the end of the study period. The control variables included race/ethnicity, age group, gender, detected infection at any time prior to vaccination (including before the study period), and the variables constructed to measure levels of non-vaccination in each staff members’ residential neighborhood and shift cohort, as described above. We also included an indicator of the shift most often worked (night, day, or swing), the number of shifts worked in the study period, and the average number of shifts worked on active weeks, as well as a prison fixed effect, indicating the prison at which a staff member worked most during the study period.

Results from the multivariable analyses are reported as the predicted probabilities that staff members in the groups of interest remained unvaccinated through June 30, 2021. For the continuous variables measuring proportion unvaccinated in residential zip codes and the shift cohorts, we report predicted values at the 25^th^, 50^th^, and 75^th^ percentile of the distributions for custody and healthcare staff, respectively. All analyses were performed using Stata version 16.1.

## Results

### Prevalence of Vaccination and Prior Infection

Uptake of Covid-19 vaccines among staff was brisk initially but plateaued within a couple of months. Figure 1 shows that from December 22, 2020 through February 15, 2021, coverage with at least 1 vaccine dose (blue and green shading) increased from 0% to 26% (0.5% per day) for custody staff and from 0% to 52% (1.0% per day) for healthcare staff. Thereafter, the pace of vaccination coverage slowed, reaching 39% (0.3% per day) for custody staff and 64% (0.5% per day) for healthcare staff by June 30, 2021. Figure 1 also shows that the surge in Covid-19 cases in the winter of 2020-21 was associated with substantial increases in the cumulative prevalence of confirmed Covid-19 cases among staff (green and yellow shading). Through December 22, 2020, CDCR had previously detected Covid-19 in 5% of the custody staff in our study sample and 2% of healthcare staff; by June 30, 2021, these proportions had increased to 27% and 12%, respectively. Seventy three percent of unvaccinated custody staff and 88% of unvaccinated healthcare staff had no recorded history of SARS-CoV-2 infection. (Appendix Figures 1 and 2 reproduce the information shown in Figure 1 for each prison).

**Figure 1:**
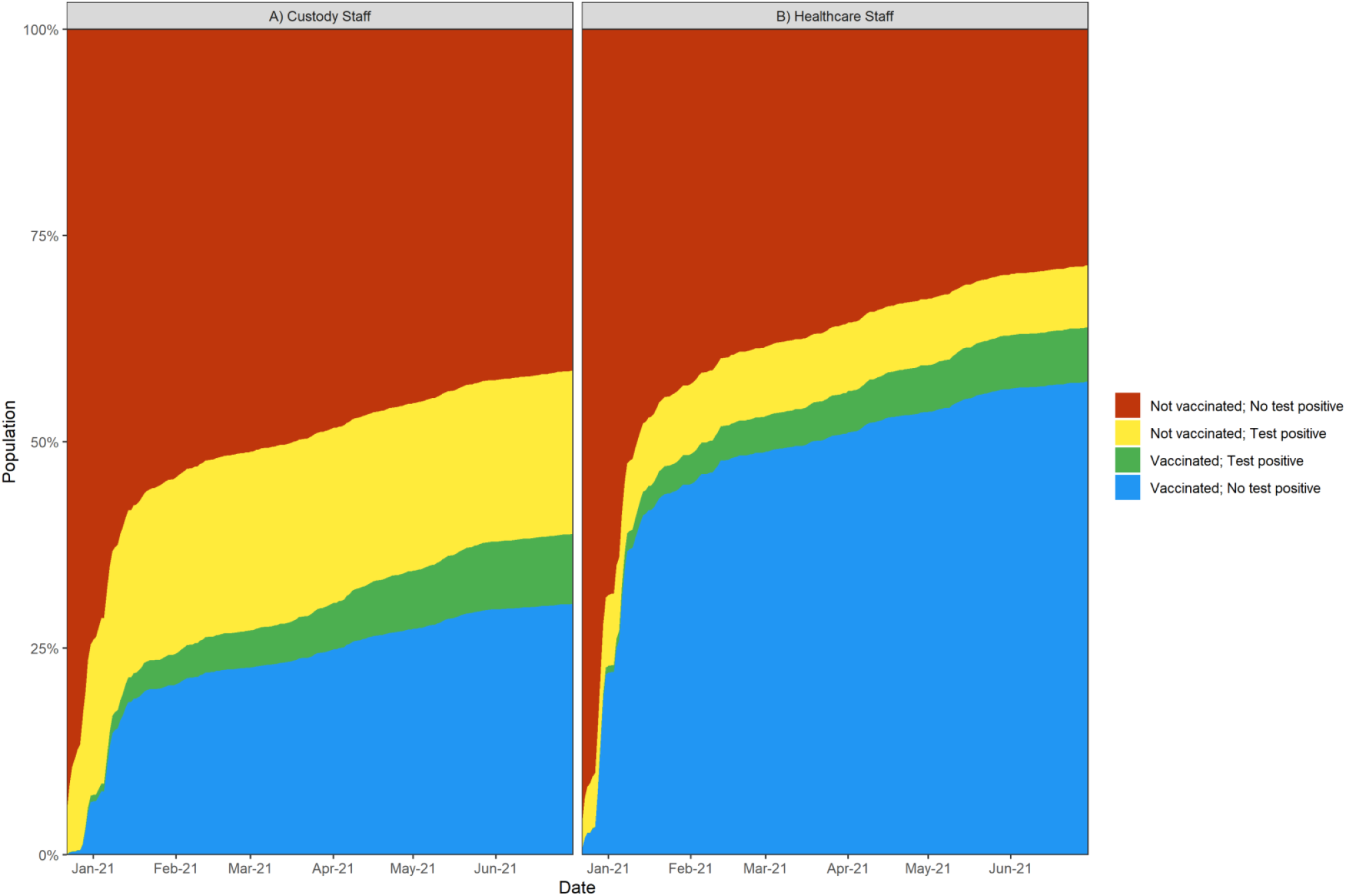
Cumulative vaccination and history of Covid-19 infection rates for custody and healthcare staff. Cumulative counts among custody and healthcare direct-care staff who had active shifts in June 2021 (N = 23,472 for custody and N = 7,617 for healthcare staff). Vaccinated indicates receipt of 1 or more doses of any Covid-19 vaccine and not vaccinated indicates receipt of zero doses of vaccine as of a given date. No test positive indicates no positive result for a recorded test for SARS-CoV-2 infection as of a given date. Test positive indicates at least one recorded positive test for infection as of a given date. Not vaccinated; no test positive represents the fraction of staff who remained unvaccinated and without a test positive through a given date. Counts do not reflect the order of vaccine uptake and SARS-CoV-2 infection, rather they shift proportionately as staff members move from category to category.

Thus, 61% of custody staff and 37% of healthcare staff remained unvaccinated at the end of the study period. These proportions varied widely across the 33 prisons in our sample—from 37% to 86% among custody staff and from 26% to 79% among healthcare staff. Figure 2 (panels A and B) plots these proportions against proportions of unvaccinated residents (details provided in Appendix Table 2). Coverage among custody staff was lower than coverage among residents in every prison; this decrement ranged from 17 to 60 percentage points. Coverage among healthcare staff was lower than coverage among residents in all but one prison, with decrements ranging from to 1 to 48 percentage points. Prisons that had higher proportions of unvaccinated custody staff also tended to have higher proportions of unvaccinated healthcare staff (Spearman’s rho: 0.57; p<0.01).

**Figure 2:**
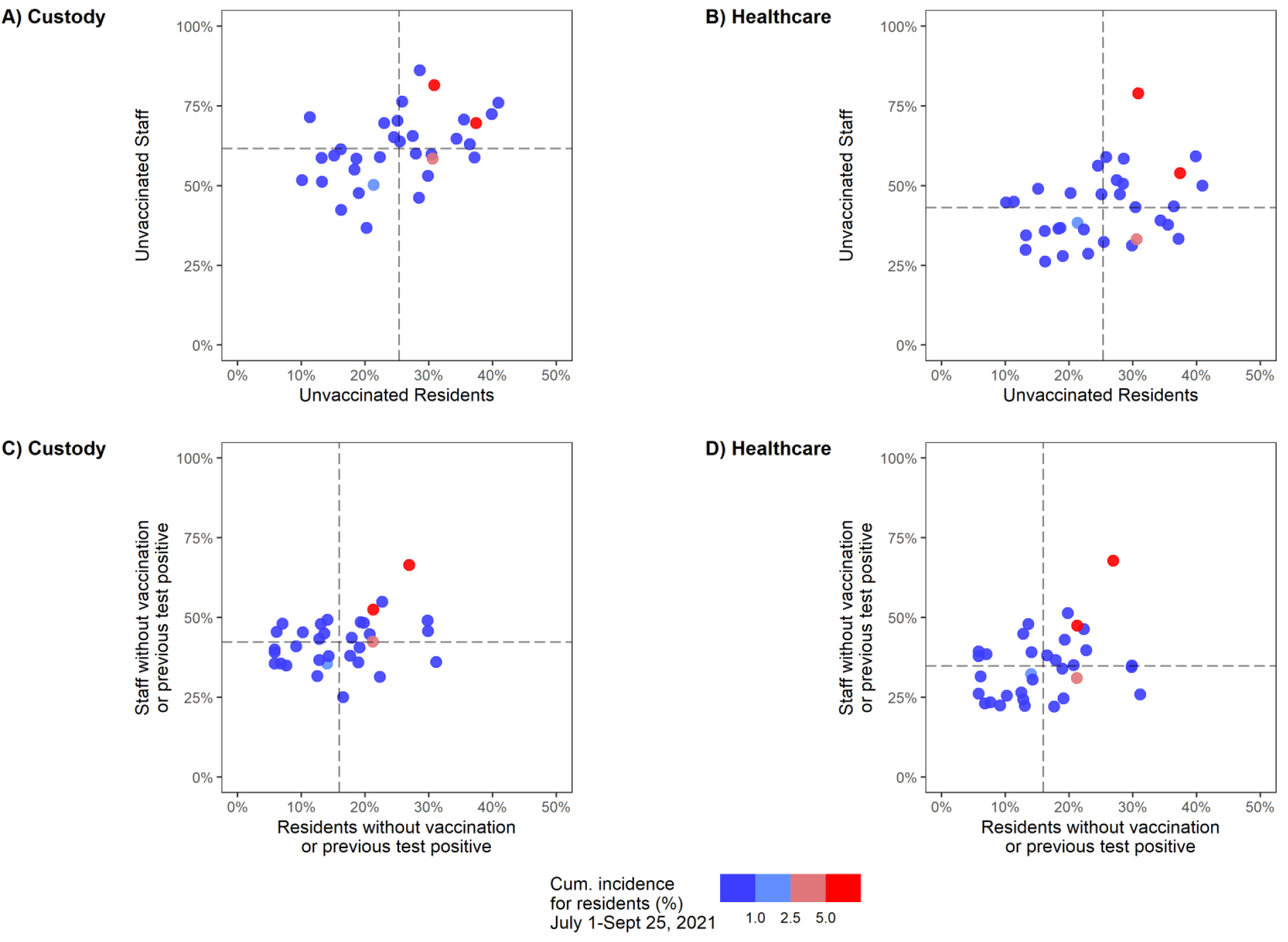
Percentage of custody staff, healthcare staff, and residents of California state prisons who are unvaccinated and have not previously tested positive for SARS-CoV-2, by prison on June 30, 2021 and cumulative incidence of infection among residents from July 1, 2021 through September 25, 2021. Proportions of unvaccinated custody and healthcare staff and residents (Panels A and B) and proportions of unvaccinated custody and healthcare staff and residents who also have no previous positive test (Panels C and D) as of June 30, 2021 at each of 33 CDCR prisons. Cumulative incidence for residents is based on total incidence of SARS-CoV-2 infection between July 1, 2021 and September 25, 2021. Vaccinated indicates receipt of 1 or more doses of any Covid-19 vaccine and not vaccinated indicates receipt of zero doses of vaccine.

The cumulative proportion of residents at each prison who became infected in the three months following the study period ranged from 0% to 7.3% (mean: 0.8%; 6 prisons above the mean) (Figure 2, Appendix Table 2). Of the 6 prisons with the largest proportions of residents infected in this 3-month period, 4 had above-average levels of unvaccinated custody staff, 4 had above-average levels of unvaccinated healthcare staff, and 5 had above-average levels of unvaccinated residents.

Patterns remained consistent when we refined the definition of the “vulnerable” population to include only individuals who remained unvaccinated and with no history of Covid-19 (Figure 2, panels C and D).

### Correctional Staff and Predictors of Remaining Unvaccinated

The study sample consisted of 31,089 prison staff employed at 33 prisons; 23,472 (75.5%) of them were custody staff and 7,617 (24.5%) were healthcare staff (Table 1). A majority were aged 30 to 49 years.Most custody staff were male (84%), and most healthcare staff were female (71%). Two thirds of custody staff were Hispanic (38%) or white (28%), while half of healthcare staff were Asian (28%) or white (23%).

**Table 1.** Individual and shift characteristics of the study sample of custody and healthcare direct-care staff at 33 California state prisons.

|  |  | Custody Staff<br>(N = 23,472)<br>n(%) |  | Health Staff<br>(N = 7,617)<br>n(%) |  |
| --- | --- | --- | --- | --- | --- |
| Individual Characteristics |  |  |  |  |  |
| Age category | 18-29 | 2,862 | (12%) | 422 | (6%) |
|  | 30-39 | 7,724 | (33%) | 2,076 | (27%) |
|  | 40-49 | 7,765 | (33%) | 2,362 | (31%) |
|  | 50-59 | 4,310 | (18%) | 1,886 | (25%) |
|  | 60+ | 811 | (3%) | 871 | (11%) |
| Race | Black | 1,571 | (7%) | 1,201 | (16%) |
|  | Hispanic | 9,008 | (38%) | 1,409 | (18%) |
|  | White | 6,666 | (28%) | 1,771 | (23%) |
|  | Other/Unknown | 4,773 | (20%) | 1,088 | (14%) |
|  | Asian/PI | 1,454 | (6%) | 2,148 | (28%) |
| Gender | Female | 3,751 | (16%) | 5,434 | (71%) |
|  | Male | 19,721 | (84%) | 2,183 | (29%) |
| No history of Covid-19 before June 30, 2021 |  | 17,052 | (73%) | 6,692 | (88%) |
| Remained unvaccinated through June 30, 2021 |  | 14,369 | (61%) | 2,822 | (37%) |
| Fraction of 20-64 year-olds vaccinated in zip code of residence (mean/SD) |  | 56% | (12%) | 52% | (13%) |
| Institution/Work Shift Characteristics |  |  |  |  |  |
| Fraction of shift co-workers not vaccinated (mean/SD) |  | 60% | (11%) | 34% | (10%) |
| Most worked shift | Night | 3,170 | (14%) | 699 | (9%) |
|  | Day | 13,510 | (58%) | 5,014 | (66%) |
|  | Swing | 6,792 | (29%) | 1,904 | (25%) |
| Number of shifts during study period (mean/SD) |  | 126 | (28) | 123 | (45) |
| Number of shifts/week (mean/SD) |  | 4.7 | (0.84) | 4.6 | (1.32) |

The analytic dataset includes 31,089 custody or healthcare staff who worked at least five direct-care shifts between April, May, and June 2021 and at least 1 direct-care shift in each of those months in any one of 33 CDCR prisons. No history of Covid-19 indicates no positive result on any PCR or antigen diagnostic test for SARS-CoV-2 infection prior to first dose of vaccination, or before the end of the study period for unvaccinated. Vaccination rates of zip code of residence are based on cumulative CDPH vaccination rates as of June 1, 2021 in the zip code where the staff person lived on the most recent recorded work shift. Fraction of co-workers not vaccinated is a count of all co-workers by date, shift, prison and is weighted by the number of shifts worked with a given co-worker. Number of shifts worked and number of shifts per week are based on shifts worked between December 22, 2020 and June 30, 2021 (for the latter, weeks in which there were zero shifts worked were excluded in calculation).

Among custody staff, 61% were unvaccinated (i.e., had received no doses of vaccine) and 72% had no confirmed Covid-19 by the end of June 2021. Among health care staff, 37% were unvaccinated and 88% had no confirmed infection. Custody staff resided in zip codes with a slightly higher rate of unvaccinated adults than healthcare staff did (56% vs. 52%, p<0.001), and had a much higher fraction of unvaccinated coworkers in their work cohorts (60% vs. 34%, p<0.001). The two types of workers had similar shift patterns.

Adjusted analyses showed a strong age gradient to vaccine uptake (Figure 3, Appendix Table 3, Appendix Table 4). For example, custody staff aged 18-29 years were 30 percentage points more likely to remain unvaccinated than their colleagues aged 60 years or older, while healthcare staff aged 18-29 years were 23 percentage points more likely to remain unvaccinated than their colleagues aged 60 years or older. Custody and healthcare staff with a history of Covid-19 were both 8 percentage points more likely to remain unvaccinated than their colleagues without such a history.

**Figure 3:**
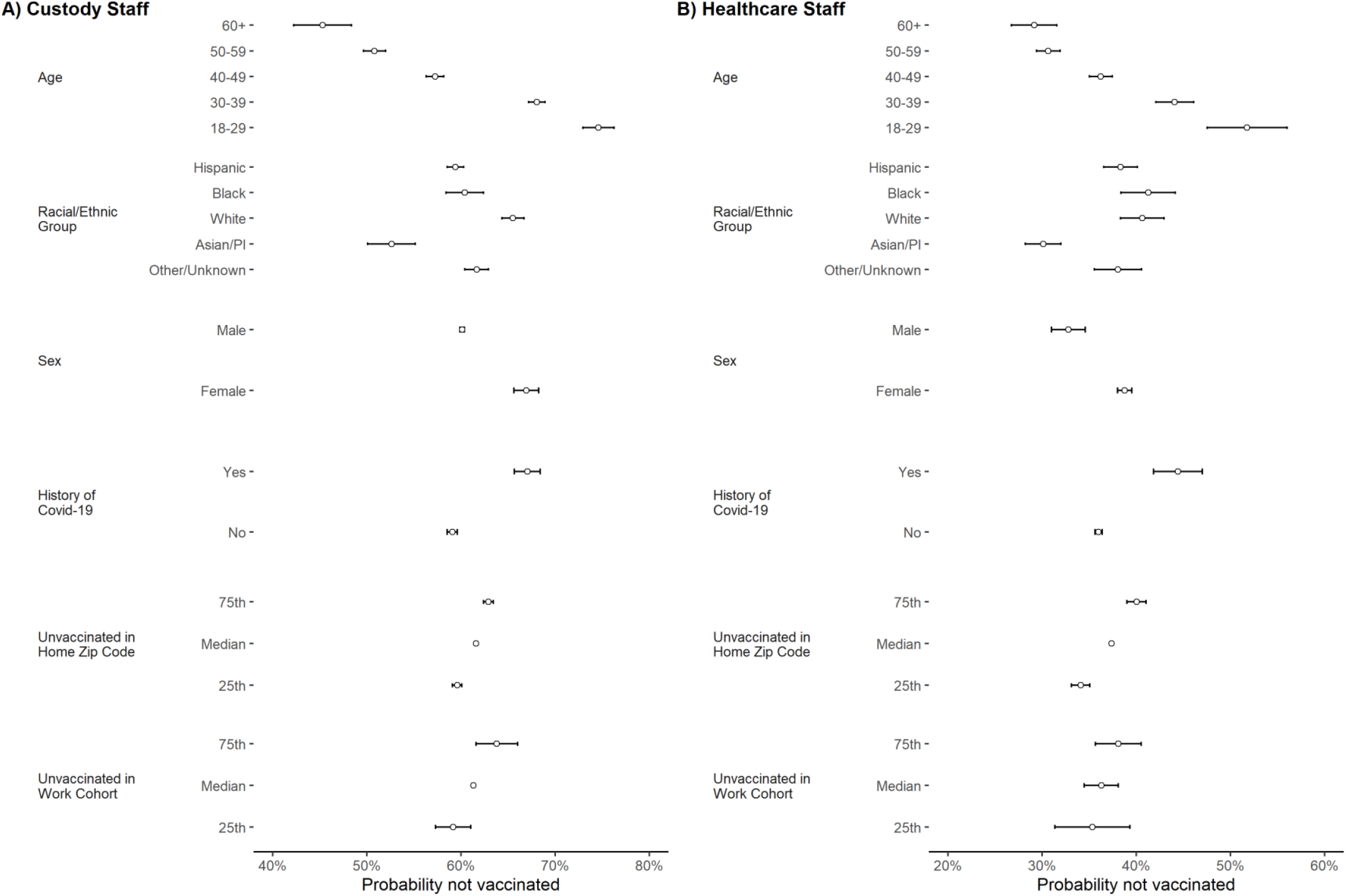
Predicted probabilities of remaining unvaccinated through June 30, 2021 for Custody and Healthcare Staff from the multivariable regression models. Panel A and Panel B show predicted probability (average adjusted margins) of being unvaccinated by June 30, 2021 for custody staff (N = 23,472) and healthcare staff (N = 7,617) respectively and was estimated using a multivariable probit model with robust standard errors, clustering on the main prison of employment. History of Covid-19 indicates a positive result on any PCR or antigen diagnostic test for SARS-CoV-2 infection during the study period. Vaccination rates of zip code of residence are based on cumulative CDPH vaccination rates as of June 1st in the zip code where the staff person lived on the most recent work-shift. Fraction of co-workers not vaccinated is a count of all co-workers by date, shift, prison and is weighted by the number of shifts worked with a given co-worker. The model controlled for all variables presented in the figure as well as an indicator of the shift (night, day, or swing) most often worked, the number of shifts worked in the study period, the average number of shifts worked on active weeks, and prison fixed effects, representing the main prison in which a staff member worked during the study period. For unvaccinated in home zip code and unvaccinated in work cohort, which are continuous variables in our model, we predict at the 25^th^, 50^th^ and 75^th^ percentiles (49%, 57%, and 63% for the former and 53%, 59%, and 67% for the latter for custody staff; 44%, 53%, and 61% for the former and 26%, 31% and 39% for the latter for healthcare staff).

Residing in zip codes with relatively large proportions of unvaccinated 18-64 year-old residents was associated with slightly higher probability of remaining unvaccinated: 3 percentage points higher for custody staff, comparing the 75^th^ percentile to the 25^th^ percentile, and 6 percentage points higher for healthcare staff (both differences significant at the p<0.05 level). Similarly, staff members who tended to share shifts with other staff that had lower levels of vaccination had a higher probability of remaining unvaccinated: 5 percentage points higher for custody staff, again comparing the 75^th^ percentile to the 25^th^ percentile, and 3 percentage points higher for healthcare staff (custody staff difference was significant at the p<0.05 level; healthcare staff difference was not). See the Supplementary Appendix for discussion of sensitivity analyses and presentation of results.

## Discussion

This study found that vaccine coverage of staff working at California state prisons lagged far behind coverage of residents for the first half of 2021. Some prisons have relatively low rates of coverage for both staff and residents, creating substantial risks of ongoing outbreaks. Our results highlight the multifactorial nature of reasons for hesitancy—a function of demographic factors, community environment, peer behaviors, and no doubt other influences that our study was not designed to measure.

Personal decisions regarding vaccination are complex.(14) Among prison staff in California we detected evidence of multi-factorial explanations. Lower uptake among younger staff and those with a history of Covid-19 are consistent with findings from other settings and suggest perceptions of one’s personal risk of infection or serious health consequences are influential.(15) The positive association we identified between low uptake and both working with unvaccinated staff and living in communities with low vaccination levels is not surprising. People are influenced by those around them, and seek out those whose beliefs align with their own. However, the risk such staff pose to residents is likely to be especially high, as they are at relatively high risk of acquiring SARS-CoV-2 infection and bringing it into the workplace. And while one might expect that in high transmission risk settings like prisons, staff would seek to protect themselves especially if those around them are less well protected via vaccination, our findings highlight the potential interplay of social dimensions and individual factors in the decision to obtain Covid-19 vaccination.

To address the risks posed by unvaccinated workers to vulnerable institutionalized populations, vaccine mandates have been enacted or proposed.(16) Although unions in these settings generally endorse vaccines, they oppose mandates and have fought them on picket-lines and in court. In California, vaccine mandates for state prison employees have come from two sources: the executive branch^6^ and the courts,(17) through an order issued in response to a request from the receiver charged with overseeing the prison health system. However, both orders met stiff opposition from the prison guards’ union and are currently mired in litigation.(18)

Our findings that correctional staff are more likely to remain unvaccinated if the people around them are unvaccinated suggests that important peer effects may be dampening vaccination uptake. Delivery of staff vaccination in the context of prisons, with or without a mandate, should be designed with peer effects in mind. For example, ensuring that individual staff members have access to vaccination in venues and at times where they can avoid being observed by their coworkers may be important. Likewise, as vaccination coverage increases in the prison and in particular staff groups (e.g., custody staff, night shift, etc.), communicating the current status of the cohort as a whole may help to shift staff member’s perception of their peers’ actions.

Our study has several limitations. First, generalizability to prisons outside California and to workers in other high-risk settings is unknown. Second, designing and implementing optimal strategies for boosting uptake of Covid-19 vaccines requires detailed understanding of the knowledge, beliefs, and preferences of the hesitant, and we did not measure these factors. Third, CDCR records may have missed vaccines obtained by some staff outside CDR’s program. However, such misclassification is likely to have been uncommon because CDCR required notification and staff who did not show proof of vaccination were required to undergo a continuous testing regimen. Fourth, our measures of co-worker and home community influences are crude; studies with qualitative and mixed-method designs are better able to engage with the nuances of personal motivations. Finally, because of the rapid rollout and sharp uptake of vaccination among staff members who chose to get vaccinated, we were unable to analyze uptake decisions over time as a function of personal experiences with infection and co-worker vaccine decisions.

Despite ongoing risks of Delta-variant Covid-19 outbreaks in high-risk carceral settings like the state prisons of California, vaccination rates among prison staff continue to lag those of incarcerated residents. Younger staff, and staff who have had Covid-19, staff who are disproportionately opting to remain unvaccinated may be doing so in part because they perceive themselves to be at low risk of infection or disease relative to their older colleagues or those who have not had a prior infection. However, social considerations both at home and at work also appear important, leading to some prisons and work cohorts with persistently below-average levels of staff vaccination. While effective and acceptable approaches to rapidly increasing vaccination coverage among correctional staff remain elusive, failing to develop and implement them prolongs very real health risks both to staff themselves, to the communities in which they reside, and to incarcerated residents.

With almost 1.2 million people currently residing in state and federal prisons in the United States, (19) addressing the risk of Covid-19 transmission and subsequent outbreaks, especially from highly transmissible viral variants like Delta, is a critical public health priority. Despite substantial levels of natural and acquired immunity in many incarcerated populations, Covid-19 risks continue to loom large, as evidenced by large summer outbreaks across California and Texas.(20, 21)

## Data Availability

The California Department of Corrections and Rehabilitation provided anonymized data which is kept on an encrypted and secure server. The data are not publicly available.

