## Supplementary Appendix for "Uptake of Covid-19 vaccines among frontline workers in California state prisons"

### Contents

Analytic cohort design ... 2

Study period ... 2

Dataset ... 2

Outcome ... 2

Variables ... 2

IRB and ethics approval ... 4

Sensitivity Analyses (SA) ... 4

Appendix Table 1: Counts of direct care staff, different zip codes of residence, and different counties of residence among CDCR custody and healthcare staff ... 6

Appendix Table 2: Fraction unvaccinated and fraction not vaccinated and without history of Covid-19) for prison for staff and incarcerated people; Cumulative incidence following the study period focused on vaccination ... 8

Appendix Table 3: Predicted probabilities and confidence intervals for adjusted analysis ... 10

Appendix Table 4: Coefficients from multivariable analyses ... 11

Appendix Table 5: Sensitivity analyses results matrix ... 12

Appendix Figure 1: Variation among prisons in vaccine and test-positive scale-up over time, Custody staff ... 13

Appendix Figure 2: Variation among prisons in vaccine and test-positive scale-up over time Healthcare staff ... 14

Analytic cohort design

Study period

The first day of the study period was the first day that vaccines were offered and recorded in the CDCR dataset, December 22, 2020. The study period goes through June 30, 2021. We chose to analyze data through June 30, 2021 for several reasons. First, we wanted to examine how vaccine uptake is correlated with subsequent outbreaks at prisons (Figure 2 in main text). Second, we are interested in mutable factors which influence vaccine uptake. After June 2021, the Delta variant became prevalent in CDCR prisons and talk of mandated vaccines for corrections workers began. Because it would be difficult to separate the difference in characteristics of late adopters from the influence of these two important changes, we chose to cut off our study period prior to July 1, 2021.

Dataset

Daily data extracts provided by the California Department of Corrections and Rehabilitation (CDCR) included unique pseudo-identifiers for both staff and incarcerated residents enabling us to follow them over time and included comprehensive information on PCR and antigen testing and vaccination. Our sample included custody and healthcare staff who worked in a CDCR prison between January 1, 2020 through June 30, 2021. The staff data were provided per day per shift, so any staff person who worked an active shift on a given day had an observation. We did not have information about leaves of absence, time off, resignation or termination other than what was imputed based on the active shift-days.

To limit our analytic dataset to correctional staff with the greatest likelihood of direct contact with incarcerated residents, we imposed several inclusion/exclusion criteria. Our analytic cohort was drawn from staff in 33 of CDCRs 35 prisons. Two of CDCR's 35 prisons were excluded from the analysis due to missing or incomplete staff data at those prisons at the time of analysis. We included only staff designated as custody or healthcare workers in one of the 33 included prisons (~63% of total CDCR staff). Other possible designations are contractor, education, and operations. Among the healthcare and custody staff, we included people designated as "direct-care" staff, meaning that they were classified by CDCR as having regular in-person contact with residents (~98% of custody and healthcare staff). We excluded staff who were missing data for any of our covariates (<1%), with the exception of missing race, which was coded under "Other/Unknown". We excluded staff who did not work at least five shifts between April 2021 and June 2021 (the period after vaccination was available to any staff person; 275 healthcare and 181 custody staff were excluded). Some prison staff (638 in our study sample) worked across multiple prisons, they entered the analysis assigned to their main institution. There were 3 staff who were appointed as custody and healthcare at different times. These were excluded from our analysis. There were 171 staff who had a record of being vaccinated in the community before their first recorded shift at CDCR. These were excluded from the analysis. Importantly, the fraction of co-workers vaccinated variable was constructed based on the entire custody or healthcare direct-care staff, prior to exclusions.

Outcome

Remained unvaccinated: CDCR staff were offered vaccines at their place of work beginning on December 22, 2020. Vaccine uptake was voluntary through the end of our study period. All vaccines administered on-site are recorded in CDCR data (and reflected in our dataset), and vaccines administered in the community were obtained from the California Immunization Registry, provided staff members gave their consent. The outcome reflects that the staff member remained unvaccinated (no doses of vaccine administered) through June 30, 2021.

Variables

*Age category:* We collapsed age into categories: 18-29, 30-39, 40-49, 50-59, 60+. These categories are consistent with or finer than those available in CDC and CDPH datasets (See, https://covid19.ca.gov/state-dashboard/#ethnicity-gender-age or https://covid.cdc.gov/covid-data-tracker/#demographicsovertime, for example).

*Race group*. We grouped race by race and ethnicity information provided by CDCR. Race categories and their components are as follows: Black (Black); Hispanic (Hispanic + Cuban + Mexican); White (White); Asian/PI (Asian or Filipino or Pacific Islander); Other/Unknown (includes American Indian or Alaskan Native (~0.4% of total sample) and other (~1% of total) and unknown or missing (~17% of total).

*Gender:* Gender is based on self-report of gender.

*History of Covid-19:* The staff data included comprehensive information on PCR and antigen testing for SARS-CoV-2 infection, with the first recorded test on March 18, 2020. Testing was voluntary and/or mandatory and occurred at varying frequency over time. Because testing was infrequent through early 2020 (see counts below), we may underestimate the proportion of staff with any history of Covid-19. Despite this, we feel confident in our estimate of fraction of staff without any positive test at the end of June 2021 because of ramped up testing starting in mid-2020 (when non-fully-vaccinated staff have been tested twice-weekly and any staff who report symptoms or work in a location with an outbreak have also been tested) in combination of the timing of most community and prison outbreaks from mid-2020 and onward.

*Monthly counts of SARS-CoV2 tests and test-positive for direct care custody and health staff*

| Month | Tests Administered | Tests Positive |
|---|---|---|
| **2020** | | |
| March | 13 | 12 |
| April | 33 | 31 |
| May | 429 | 66 |
| June | 9,729 | 215 |
| July | 66,684 | 831 |
| August | 55,047 | 535 |
| September | 72,164 | 290 |
| October | 81,580 | 382 |
| November | 97,484 | 1,163 |
| December | 121,075 | 3,272 |
| **2021** | | |
| January | 109,535 | 1,400 |
| February | 102,838 | 356 |
| March | 124,159 | 293 |
| April | 128,069 | 230 |
| May | 87,398 | 169 |
| June | 93,663 | 205 |

*Counts are for the 33 prisons included in the analytic sample.

*Unvaccinated in zip code:* Fraction of unvaccinated adults in staff home-zip code is based on the cumulative percentage of the 20-64 year-old population in a given zip-code who received 1+ doses of vaccination by June 1, 2020. Data are from CDPH (see reference in main text). We used the last known zip code of staff members to create this measure.

*Unvaccinated in work cohort:* To build a cross-sectional measure of peer vaccine take-up, we counted, for each staff-shift, all co-workers on each prison-shift-day and all coworkers who were unvaccinated on each prison-shift-day. The individual-level measure was $\frac{\sum_{S=1}^{N} Us-1}{\sum_{S=1}^{N} Ts-1}$ where *N* is the total number of shifts worked by an individual staff person during the study period, *Us* is the number of unvaccinated workers on a particular prison-shift-day and *Ts* is the total number of workers on a particular prison-shift-day. Note, we subtract the individual for whom the measure is calculated in the counts of unvaccinated and of total workers to avoid a reflection issue.

See Appendix Table 5 for a sensitivity analysis limiting the analysis to prisons with the greatest within-institution inter-quartile range (as a measure of spread).

*Shift variables:* We created a categorical variable denoting which shift a staff person worked most often (based on raw count of shifts worked over the study period): day, night, or swing shift. We controlled for the total number of shifts worked during the study period, the mean shifts per week worked (weeks with zero shifts were not counted in the mean), and their interaction.

*Prison Fixed effects:* We included prison-level fixed effects to control for stable heterogeneity between prisons.

IRB and ethics approval

The study was approved by the institutional review board (IRB) at Stanford University (protocol numbers IRB-55835, IRB-55671). The IRB approval of the study included a waiver of consent, on the basis that CDCR provided the Stanford research team with a limited data set without direct identifiers, the data had been collected for operational purposes, and the study could not practicably be carried out otherwise. Similar approval conditions were met for California Department of Public Health data.

Sensitivity Analyses (SA)

We ran three sensitivity analyses to explore questions related to the fraction of co-workers unvaccinated and the history of Covid-19 variables, as follows. These were run on the custody staff group. Predicted probabilities and 95% confidence intervals are presented in Appendix Table 5.

*SA 1: Within prison dispersion in the fraction of co-workers vaccinated*

As noted in the text, our measure of co-worker peer influence is crude. To estimate the relationship between co-worker vaccination uptake and individual staff decision to remain unvaccinated, we would ideally see substantial variation across individual staff workers in the rate of vaccination of their co-worker cohorts within a given prison – a signal that there are differences in cohort-preferences for vaccination. If there is not substantial variation, it could be possible that exogenous shocks over time (not accounted for by the prison fixed effects) lead to heterogeneity in cohort vaccination rates. (Note: because the patterns of vaccination uptake are strikingly similar across prisons [see Appendix Figures 1 and 2] this concern about variation in the timing of vaccination due to such shocks is substantially reduced.) To explore this further, we examined the within-prison dispersion in the fraction of (custody staff) co-worker cohorts vaccinated and repeated our main regression analysis on the sample after excluding those prisons with the least variation. Within prisons, the mean fraction of unvaccinated custody workers was between 34% and 86%, the range (highest value – lowest value) was between 4 percentage points and 24 percentage points, and the interquartile range (IQR) between 1 and 8 percentage points. We re-ran our multivariable analysis limiting the sample to those prisons with a wider distribution of values (IQR ≥ median; 17 of 33 prisons included; N = 12,221) to examine whether individuals working in prisons with a wider dispersion were more or less likely to remain unvaccinated if their co-workers we less likely to be vaccinated. The results were robust to our main analysis, custody staff were six percentage points more likely to remain unvaccinated if their co-worker cohort was at the 75$^{th}$ percentile of being unvaccinated compared to working with a cohort at the 25$^{th}$ percentile (compared to being 5 percentage points more likely in our main analysis). See Appendix Table 5 for results.

*SA 2: Interaction of main shift worked with fraction of co-workers vaccinated*

It may be the case that peer interactions may be different for night shift versus day or swing shift and hence the relationship between co-worker vaccination patterns and staff remaining unvaccinated may be different by shift. To assess the possibility that main-shift-worked is an effect modifier (e.g., the correct model specification interacts the shift-variable with the fraction of co-worker vaccinated variable) in our model, we repeated the multivariable probit regression as in the main analysis expanded to include this interaction term. The coefficient on the interaction term was not significantly different than zero and the predicted probabilities were consistent with those resulting from our main specification. See Appendix Table 5 for results.

*SA 3: History of Covid-19 by time of test*

Because there is some concern that vaccination uptake is related to the timing of a previously positive SARS-CoV-2 test (e.g., an individual may be advised to wait 90 days from last positive test to receive a dose of vaccine), we divided our *History of Covid-19* variable into time periods as follows. The reference group (as in the main specification) is “no history of Covid-19” (N = 17,052), and the other categories are: last positive test prior to December 22, 2020 (N = 4,619); last positive test between December 22, 2020 and March 15, 22021 (N = 1,423); last positive test after March 15, 2021 (N = 378). We chose the time periods based on the idea that all those in the first and second groups (e.g., before March 15, 2021) would have time to become vaccinated within our study period even after a 90-day post infection window, and those in the first group would have time to be become vaccinated during the initial vaccine push evident in Figure 1 in the main text. We find that, for residents with last positive SARS-CoV-2 test prior to December 22, they are 5 percentage points more likely to remain unvaccinated compared to those with no prior history of Covid-19. For those with last positive SARS-CoV-2 test in the first few months of the study period, they are 13 percentage points more likely to remain unvaccinated. We can compare this to an estimate of those with prior infection in the main specification being 8-percentage points more likely to remain unvaccinated than those with no prior infection. Thus, when focusing on those with a history of prior Covid-19 more than 90 days before the end of our study period, history of prior Covid-19 remains a significant predictor of remaining unvaccinated.

Appendix Table 1: Counts of direct care staff, different zip codes of residence, and different counties of residence among CDCR custody and healthcare staff

| | Custody staff | | | Healthcare Staff | | |
|---|---|---|---|---|---|---|
| Prison | # Direct Care Staff | # of Unique Zip Codes of Residence | # of Counties of Residence | # Direct Care Staff | # of Unique Zip Codes of Residence | # of Unique Counties of Residence |
| 1 | 675 | 72 | 9 | 96 | 32 | 8 |
| 2 | 358 | 75 | 8 | 64 | 17 | 4 |
| 3 | 687 | 37 | 6 | 97 | 30 | 8 |
| 4 | 502 | 81 | 30 | 59 | 11 | 10 |
| 5 | 921 | 63 | 11 | 151 | 40 | 8 |
| 6 | 524 | 69 | 12 | 213 | 50 | 13 |
| 7 | 656 | 35 | 6 | 101 | 31 | 7 |
| 8 | 1027 | 120 | 18 | 1518 | 177 | 31 |
| 9 | 805 | 179 | 8 | 267 | 112 | 12 |
| 10 | 457 | 147 | 8 | 289 | 121 | 6 |
| 11 | 799 | 42 | 13 | 271 | 34 | 11 |
| 12 | 822 | 115 | 20 | 641 | 125 | 28 |
| 13 | 1151 | 75 | 9 | 316 | 69 | 17 |
| 14 | 736 | 149 | 7 | 115 | 64 | 5 |
| 15 | 687 | 69 | 21 | 133 | 46 | 17 |
| 16 | 382 | 81 | 8 | 60 | 23 | 6 |
| 17 | 460 | 88 | 15 | 115 | 50 | 12 |
| 18 | 530 | 87 | 13 | 110 | 42 | 7 |
| 19 | 626 | 63 | 23 | 110 | 27 | 17 |
| 20 | 570 | 104 | 7 | 76 | 28 | 7 |
| 21 | 912 | 52 | 6 | 183 | 45 | 11 |
| 22 | 762 | 156 | 11 | 257 | 91 | 10 |
| 23 | 810 | 103 | 15 | 298 | 96 | 18 |
| 24 | 540 | 23 | 15 | 65 | 15 | 12 |
| 25 | 752 | 66 | 7 | 115 | 24 | 6 |
| 26 | 938 | 136 | 14 | 347 | 96 | 10 |
| 27 | 854 | 114 | 19 | 318 | 85 | 19 |
| 28 | 938 | 67 | 8 | 292 | 70 | 19 |
| 29 | 576 | 155 | 26 | 70 | 31 | 8 |

| | | | | | | |
|---|---|---|---|---|---|---|
| 30 | 666 | 106 | 21 | 137 | 54 | 11 |
| 31 | 991 | 254 | 35 | 256 | 105 | 15 |
| 32 | 868 | 127 | 26 | 299 | 101 | 23 |
| 33 | 490 | 63 | 9 | 178 | 44 | 10 |

All counts are derived from the analytic sample used in the multivariable analysis.

Appendix Table 2: Fraction unvaccinated and fraction not vaccinated and without history of Covid-19) for prison for staff and incarcerated people; Cumulative incidence following the study period focused on vaccination

| | Incarcerated persons | | Custody Staff | | Healthcare staff | | Cumulative Incidence (July 1 - Sept 25, 2021) |
|---|---|---|---|---|---|---|---|
| Prison | Not vaccinated no history of Covid-19 (%) | Not vaccinated (%) | Not vaccinated no history of Covid-19 (%) | Not vaccinated (%) | Not vaccinated no history of Covid-19 (%) | Not vaccinated (%) | Fraction of Incarcerated Persons (%) |
| 1 | 6% | 15% | 39% | 59% | 39% | 49% | 0.12% |
| 2 | 19% | 36% | 41% | 71% | 25% | 38% | 0.58% |
| 3 | 22% | 28% | 31% | 46% | 46% | 51% | 0.03% |
| 4 | 23% | 41% | 55% | 76% | 40% | 50% | 0.99% |
| 5 | 19% | 26% | 49% | 76% | 43% | 59% | 0.31% |
| 6 | 21% | 28% | 45% | 60% | 35% | 47% | 0.37% |
| 7 | 17% | 20% | 25% | 37% | 38% | 48% | 0.11% |
| 8 | 10% | 13% | 45% | 59% | 25% | 30% | 0.48% |
| 9 | 13% | 18% | 32% | 55% | 26% | 36% | 0.21% |
| 10 | 14% | 21% | 35% | 50% | 32% | 38% | 1.21% |
| 11 | 8% | 25% | 35% | 64% | 23% | 32% | 0.26% |
| 12 | 13% | 19% | 37% | 48% | 24% | 28% | 0.40% |
| 13 | 18% | 25% | 44% | 70% | 37% | 47% | 0.49% |
| 14 | 9% | 22% | 41% | 59% | 22% | 36% | 0.81% |
| 15 | 6% | 10% | 40% | 52% | 38% | 45% | 0.04% |
| 16 | 6% | 16% | 45% | 61% | 31% | 36% | 0.05% |
| 17 | 13% | 23% | 48% | 70% | 22% | 29% | 0.00% |
| 18 | 18% | 30% | 38% | 53% | 22% | 31% | 0.04% |
| 19 | 14% | 29% | 45% | 86% | 48% | 58% | 0.58% |
| 20 | 14% | 28% | 49% | 66% | 39% | 52% | 0.00% |
| 21 | 30% | 36% | 46% | 63% | 35% | 43% | 0.03% |
| 22 | 19% | 30% | 36% | 60% | 34% | 43% | 0.22% |
| 23 | 7% | 11% | 48% | 71% | 38% | 45% | 0.46% |
| 24 | 27% | 31% | 66% | 82% | 68% | 79% | 6.22% |
| 25 | 20% | 40% | 48% | 72% | 51% | 59% | 0.94% |
| 26 | 14% | 19% | 38% | 58% | 31% | 37% | 0.15% |

| | | | | | | | |
|---|---|---|---|---|---|---|---|
| 27 | 30% | 34% | 49% | 65% | 34% | 39% | 0.71% |
| 28 | 13% | 25% | 43% | 65% | 45% | 56% | 0.32% |
| 29 | 21% | 37% | 52% | 70% | 47% | 54% | 7.29% |
| 30 | 21% | 31% | 42% | 58% | 31% | 33% | 3.38% |
| 31 | 7% | 16% | 36% | 42% | 23% | 26% | 0.28% |
| 32 | 31% | 37% | 36% | 59% | 26% | 33% | 0.00% |
| 33 | 6% | 13% | 36% | 51% | 26% | 34% | 0.44% |

Rates are based on our study sample (see main text for details). Cumulative incidence is the rate of new detected Covid-19 cases in incarcerated persons after the study period (from July 1, 2021 - September 25, 2021).

Appendix Table 3: Predicted probabilities and confidence intervals for adjusted analysis

| | Custody staff | | Healthcare staff | |
|---|---|---|---|---|
| | Probability | 95% Confidence Interval | Probability | 95% Confidence Interval |
| **Age** | | | | |
| 60+ | 0.45 | (0.42 - 0.48) | 0.29 | (0.27 - 0.32) |
| 50-59 | 0.51 | (0.50 - 0.52) | 0.31 | (0.29 - 0.32) |
| 40-49 | 0.57 | (0.56 - 0.58) | 0.36 | (0.35 - 0.37) |
| 30-39 | 0.68 | (0.67 - 0.69) | 0.44 | (0.42 - 0.46) |
| 18-29 | 0.75 | (0.73 - 0.76) | 0.52 | (0.48 - 0.56) |
| **Racial/Ethnic Group** | | | | |
| Hispanic | 0.59 | (0.59 - 0.60) | 0.38 | (0.37 - 0.40) |
| Black | 0.6 | (0.58 - 0.62) | 0.41 | (0.38 - 0.44) |
| White | 0.66 | (0.64 - 0.67) | 0.41 | (0.38 - 0.43) |
| Asian/PI | 0.53 | (0.50 - 0.55) | 0.3 | (0.28 - 0.32) |
| Other/Unknown | 0.62 | (0.60 - 0.63) | 0.38 | (0.36 - 0.41) |
| **Gender** | | | | |
| Male | 0.6 | (0.60 - 0.60) | 0.33 | (0.31 - 0.35) |
| Female | 0.67 | (0.66 - 0.68) | 0.39 | (0.38 - 0.40) |
| **History of Covid-19 (before vaccination)** | | | | |
| Yes | 0.67 | (0.66 - 0.68) | 0.44 | (0.42 - 0.47) |
| No | 0.59 | (0.59 - 0.60) | 0.36 | (0.36 - 0.36) |
| **Unvaccinated in Home Zip Code** | | | | |
| 75th% | 0.63 | (0.62 - 0.63) | 0.4 | (0.39 - 0.41) |
| Median | 0.62 | (0.61 - 0.62) | 0.37 | (0.37 - 0.37) |
| 25th% | 0.6 | (0.59 - 0.60) | 0.34 | (0.33 - 0.35) |
| **Unvaccinated in Work Cohort** | | | | |
| 75th% | 0.64 | (0.62 - 0.66) | 0.38 | ( 0.36 - 0.41) |
| Median | 0.61 | (0.61 - 0.61) | 0.36 | ( 0.34 - 0.38) |
| 25th% | 0.59 | (0.57 - 0.61) | 0.35 | ( 0.31 - 0.39) |

See legend notes for Figure 3 in main text.

Appendix Table 4: Coefficients from multivariable analyses

| | Custody staff | Healthcare staff |
|---|---|---|
| Age (compared to 20-29) | | |
| 30-39 | -0.204*** | -0.212*** |
| 40-49 | -0.508*** | -0.436*** |
| 50-59 | -0.680*** | -0.605*** |
| 60+ | -0.827*** | -0.653*** |
| Race/Ethnicity (compared to Black) | | |
| Hispanic | -0.028 | -0.084* |
| White | 0.147*** | -0.018 |
| Other/unknown | 0.036 | -0.093 |
| Asian/Pacific Islander | -0.215*** | -0.334*** |
| Gender (Compared to Female) | | |
| Male | -0.198*** | -0.179*** |
| History of Covid-19 (before vaccination) | 0.230*** | 0.245*** |
| Fraction of adults in zip code unvaccinated | 0.669*** | 1.025*** |
| Fraction of shift co-workers unvaccinated | 0.904** | 0.605 |
| Main Shift worked (compared to night shift) | | |
| Day shift | 0.012 | -0.083* |
| Swing shift | 0.017 | 0.03 |
| Number of shifts worked | -0.007*** | -0.016*** |
| Mean # shifts per week worked | 0.127*** | 0.325*** |
| Constant | 0.138 | -0.05 |
| Observations | 23,472 | 7,617 |
| Prison Fixed Effects Included | Yes | Yes |

*** p<0.01, ** p<0.05, * p<0.1

Appendix Table 5: Sensitivity analyses results matrix

| | Main analysis (custody staff) | | SA1: limited to prisons with more variation in fraction of co-workers not vaccinated | | SA2: Interact shift most worked with unvaccinated in work cohort variable | | SA3: History of Covid-19 by time period | |
|---|---|---|---|---|---|---|---|---|
| | Probability | 95% Confidence Interval | Probability | 95% Confidence Interval | Probability | 95% Confidence Interval | Probability | 95% Confidence Interval |
| **Age** | | | | | | | | |
| 60+ | 0.45 | ( 0.42 - 0.48) | 0.43 | ( 0.39 - 0.47) | 0.45 | ( 0.42 - 0.48) | 0.45 | ( 0.42 - 0.48) |
| 50-59 | 0.51 | ( 0.50 - 0.52) | 0.47 | ( 0.45 - 0.49) | 0.51 | ( 0.50 - 0.52) | 0.51 | ( 0.50 - 0.52) |
| 40-49 | 0.57 | ( 0.56 - 0.58) | 0.53 | ( 0.52 - 0.55) | 0.57 | ( 0.56 - 0.58) | 0.57 | ( 0.56 - 0.58) |
| 30-39 | 0.68 | ( 0.67 - 0.69) | 0.65 | ( 0.64 - 0.66) | 0.68 | ( 0.67 - 0.69) | 0.68 | ( 0.67 - 0.69) |
| 18-29 | 0.75 | ( 0.73 - 0.76) | 0.73 | ( 0.71 - 0.76) | 0.75 | ( 0.73 - 0.76) | 0.75 | ( 0.73 - 0.76) |
| **Racial/Ethnic Group** | | | | | | | | |
| Hispanic | 0.59 | ( 0.59 - 0.60) | 0.57 | ( 0.55 - 0.58) | 0.59 | ( 0.59 - 0.60) | 0.59 | ( 0.58 - 0.60) |
| Black | 0.60 | ( 0.58 - 0.62) | 0.59 | ( 0.57 - 0.61) | 0.60 | ( 0.58 - 0.62) | 0.60 | ( 0.58 - 0.62) |
| White | 0.66 | ( 0.64 - 0.67) | 0.62 | ( 0.61 - 0.64) | 0.65 | ( 0.64 - 0.67) | 0.65 | ( 0.64 - 0.67) |
| Asian/PI | 0.53 | ( 0.50 - 0.55) | 0.48 | ( 0.44 - 0.51) | 0.53 | ( 0.50 - 0.55) | 0.53 | ( 0.50 - 0.55) |
| Other/Unknown | 0.62 | ( 0.60 - 0.63) | 0.58 | ( 0.57 - 0.60) | 0.62 | ( 0.60 - 0.63) | 0.62 | ( 0.60 - 0.63) |
| **Gender** | | | | | | | | |
| Male | 0.60 | ( 0.60 - 0.60) | 0.57 | ( 0.57 - 0.58) | 0.60 | ( 0.60 - 0.60) | 0.60 | ( 0.60 - 0.60) |
| Female | 0.67 | ( 0.66 - 0.68) | 0.62 | ( 0.60 - 0.64) | 0.67 | ( 0.66 - 0.68) | 0.67 | ( 0.66 - 0.68) |
| **History of Covid-19 (before vaccination)** | | | | | | | | |
| Yes | 0.67 | ( 0.66 - 0.68) | 0.63 | ( 0.61 - 0.66) | 0.67 | ( 0.66 - 0.68) | | |
| No | 0.59 | ( 0.59 - 0.60) | 0.56 | ( 0.55 - 0.57) | 0.59 | ( 0.59 - 0.60) | | |
| **History of Covid-19, by date of last positive test before vaccination** | | | | | | | | |
| No history | | | | | | | 0.59 | ( 0.59 - 0.60) |
| Before Dec 22, 2020 | | | | | | | 0.64 | ( 0.63 - 0.65) |
| Dec 22, 2020 to March 15, 2021 | | | | | | | 0.72 | ( 0.70 - 0.75) |
| After March 15, 2021 | | | | | | | 0.85 | ( 0.80 - 0.89) |
| **Unvaccinated in Home Zip Code** | | | | | | | | |
| 75th% | 0.63 | ( 0.62 - 0.63) | 0.60 | ( 0.59 - 0.61) | 0.63 | ( 0.62 - 0.63) | 0.63 | ( 0.62 - 0.63) |
| Median | 0.62 | ( 0.61 - 0.62) | 0.58 | ( 0.58 - 0.59) | 0.62 | ( 0.61 - 0.62) | 0.62 | ( 0.61 - 0.62) |
| 25th% | 0.60 | ( 0.59 - 0.60) | 0.57 | ( 0.56 - 0.57) | 0.60 | ( 0.59 - 0.60) | 0.60 | ( 0.59 - 0.60) |
| **Unvaccinated in Work Cohort** | | | | | | | | |
| 75th% | 0.64 | ( 0.62 - 0.66) | 0.61 | ( 0.59 - 0.64) | 0.64 | ( 0.62 - 0.66) | 0.64 | ( 0.62 - 0.66) |
| Median | 0.61 | ( 0.61 - 0.61) | 0.59 | ( 0.58 - 0.59) | 0.61 | ( 0.61 - 0.61) | 0.61 | ( 0.61 - 0.61) |
| 25th% | 0.59 | ( 0.57 - 0.61) | 0.55 | ( 0.54 - 0.57) | 0.59 | ( 0.57 - 0.61) | 0.59 | ( 0.57 - 0.61) |

See legend notes for Figure 3 in main text.

Appendix Figure 1: Variation among prisons in vaccine and test-positive scale-up over time, Custody staff

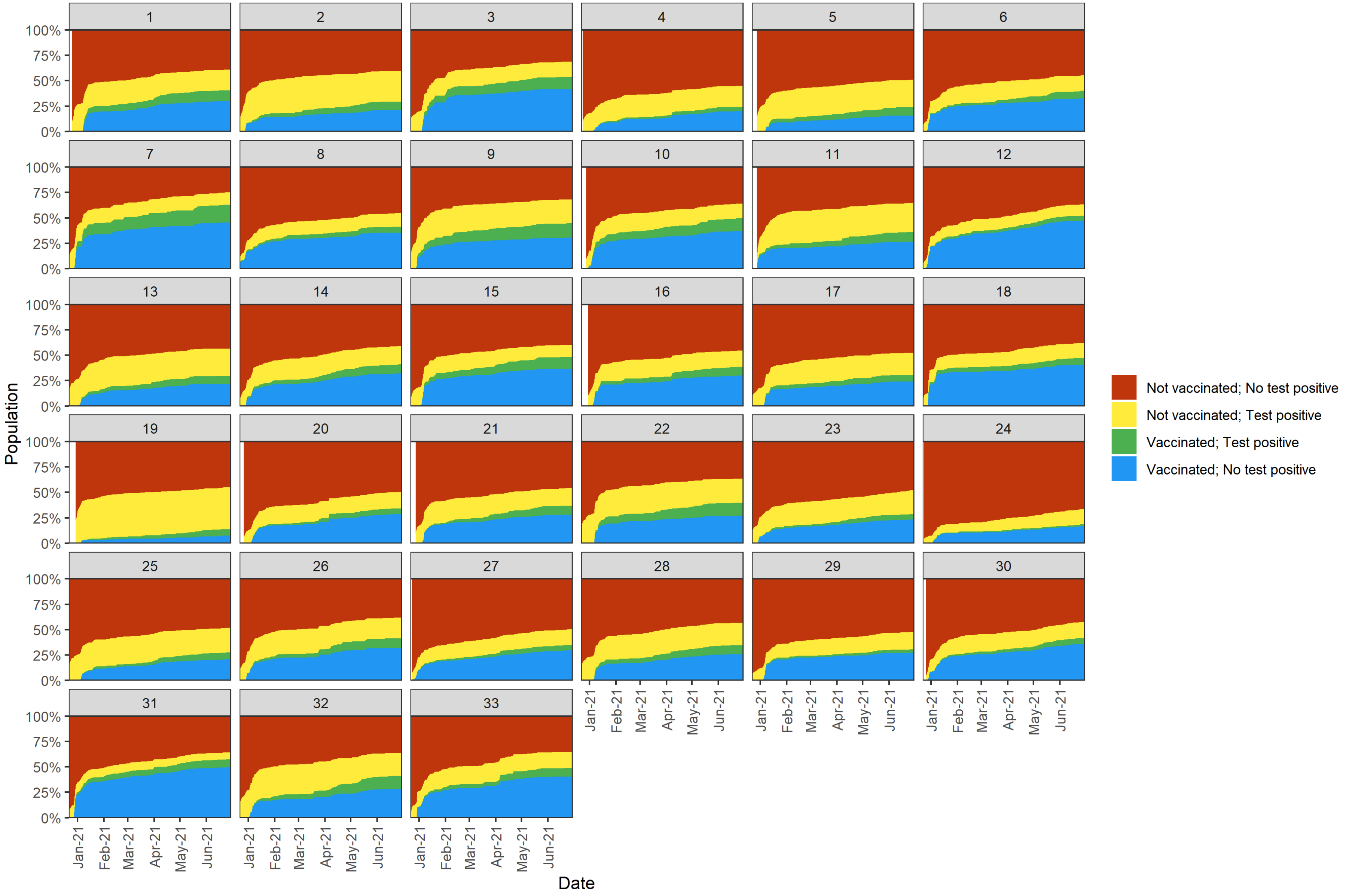

Appendix Figure 2: Variation among prisons in vaccine and test-positive scale-up over time Healthcare staff

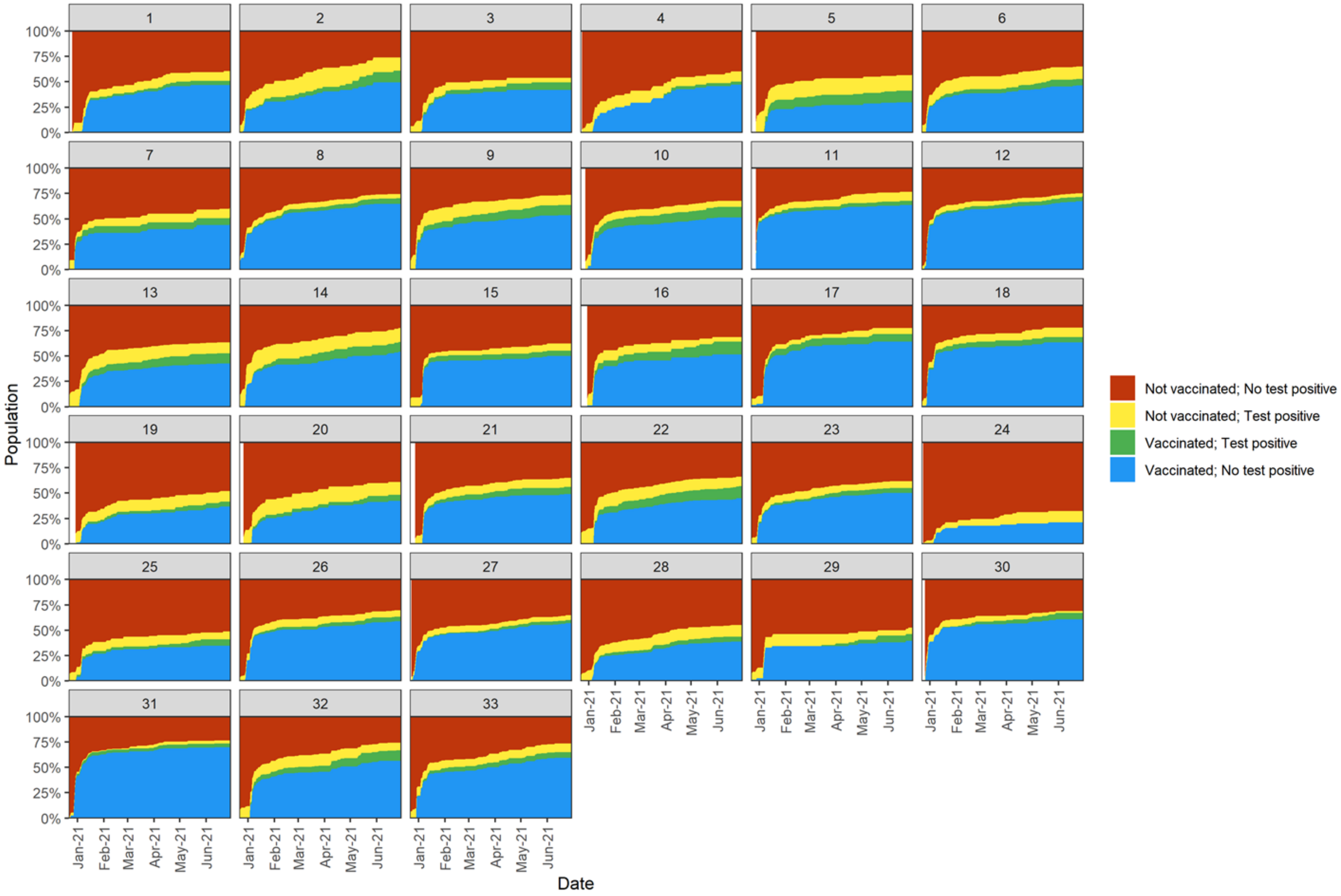
